# Socioeconomic status is not associated with survival in patients with brain metastases treated with stereotactic radiotherapy

**DOI:** 10.1101/2022.10.03.22280630

**Authors:** S.H.J. Nagtegaal, S.G. Elias, T.J. Snijders, H.M. Verkooijen, J.J.C. Verhoeff

## Abstract

**Background and purpose:** In most cancer sites a low socioeconomic status (SES) is consistently associated with poorer survival. For brain metastasis, this relation is not well understood. Therefore, we studied the effect of SES on survival in Dutch brain metastasis patients treated with stereotactic radiosurgery in a tertiary radiotherapy facility.

**Materials and methods:** We retrospectively studied 404 consecutive patients treated with stereotactic radiosurgery for brain metastases in a tertiary referral centre between 2012 and 2017. Baseline prognostic factors for survival were collected. The SES score was based on education, income and employment. Cox proportional hazard models were made, corrected for the relevant variables identified from a directed acyclic graph (DAG). Adjustments were made in two ways in order to obtain comprehensive results: correcting for confounders (total effect), and correcting for confounders and mediating factors (direct effect).

**Results:** Unadjusted, estimation of the linear effect of SES on survival resulted in a HR of 0.92 (95%CI 0.82 – 1.04). The total effect of SES was achieved by correcting for age, resulting in a HR of 0.95, with a 95% CI of 0.84 – 1.07. The direct effect gave a HR of 0.96 (95%CI 0.84 – 1.10).

**Conclusion:** SES was not associated with survival in patients with brain metastases undergoing radiation therapy. When correcting for clinical variables, we found no significant relationship between SES and survival, with HRs suggesting limited clinical impact. The results suggest that patients’ survival outcomes after contemporary cancer treatment are unrelated to their employment status, education status and annual income.

## 1. Introduction

In patients with brain metastases median survival after diagnosis ranges from 2 to 12 months, depending on the primary tumour site [1,2]. Brain metastases occur in 20-40% of all cancer patients, with 8-10% having intracranial metastasis at initial presentation [1–5]. The occurrence of brain metastases has a negative impact on survival of cancer patients, as well as affecting quality of life negatively. Treatment options for brain metastases are surgery, systemic treatment, and radiotherapy (RT), of which there are two main types: whole-brain radiotherapy and stereotactic radiotherapy (SRT). The latter can be delivered fractionated (FSRT) or in a single dose, referred to as stereotactic radiosurgery (SRS). New therapies are constantly being developed in order to improve survival after treatment. Besides therapy, other factors can have an effect on a patients’ life expectancy as well. One of these is the socioeconomic status (SES). Several reviews have shown that in most major cancer sites a low SES is consistently associated with poorer survival compared with a high SES [6–9]. These studies and reviews analysed patients from several countries in North America, Europe and Asia, with varying healthcare systems (single and multi-payer, single and multiple provider).

Factors that contribute to a patient’s SES, such as education level, income, and health insurance coverage, each influence access to healthcare, which in turn causes disparities in the rate of early detection, preventative medicine, adequate treatment and palliative care [9]. Additionally, lifestyle factors like healthy diet, smoking, alcohol consumption and regularity of exercise are correlated with SES. As with many sociological phenomena, the general effect of SES on health is complexly interlinked with other cultural factors, including attitudes towards health, belief in modern medicine, and racial bias [10,11].

Only a handful of papers have examined SES as a prognostic factor for survival in patients with brain metastases, with varying results. Most find no significant relation between SES and survival [12,13], while one Dutch and one Chinese study did see a significant effect [14,15].

The Dutch study, recently conducted by Ten Berge et al., studied a cohort of 1129 Dutch patients with synchronously diagnosed brain metastases and non-small-cell lung carcinoma (NSCLC) treated with Gamma Knife radiosurgery, and analysed whether SES influenced referral patterns and survival. Compared with low SES, patients with intermediate and high SES had a more favourable survival, with HRs of 0.9 and 0.8.

We conducted a similar analysis to Ten Berge et al. in a cohort more representative of the brain metastasis population. We selected patients referred to a tertiary radiotherapy facility in the Netherlands for linac-based radiosurgery of both synchronous and non-synchronous brain metastases from multiple primary tumour sites.

## 2. Methods

### 2.1 Participants and data collection

We identified all consecutive patients who were referred to the radiation oncology department of the UMC Utrecht for first SRS or FSRT treatment of newly diagnosed brain metastases (both synchronous and non-synchronous) between 1-1-2012 and 31-12-2017. Patients were eligible for inclusion if they had a primary tumour included in the disease-specific Graded Prognostic Assessment (DS-GPA) model, i.e. NSCLC, breast cancer, melanoma, renal cell cancer (RCC), and gastro-intestinal (GI) tumours.

We collected information on the following prognostic factors of the DS-GPA at baseline from electronic patient records: Karnofsky performance status (KPS), age, number of brain metastases, presence of extra-cranial metastases and disease-specific tumour markers. The following markers were collected: BRAF gene status (for melanoma), EGFR/ ALK gene status (for NSCLC), and HER2, and ER/PR receptor status (for breast cancer). Additionally, Hb (converted from mmol/l to g/dL) was collected for RCC from standardized laboratory examination of blood samples.

The determinant SES was determined at baseline. The Netherlands Institute for Social Research (In Dutch: *Sociaal en Cultureel Planbureau, SCP*) periodically collects data on SES per postal code [16]. For each postal code, they summarize the SES in a score, based on education level, income and employment status. The score is based on a principal component analysis and is standardised, with a national average over time of 0, and a standard deviation of 1. This means each postal code receives a single score which has no intrinsic value, but is designed to allow comparisons with other regions over time.

The primary outcome of the study was overall survival in months after the start of RT. Data on survival was retrieved from patient records and the Municipal Personal Records Database (*Gemeentelijke Basisadministratie, GBA*). Patients who were still alive were censored on the date the GBA was consulted.

The need for informed consent was waived. We obtained permission from the Ethics Board of our institution to conduct this study with reference number 18/273. To ensure protection of private data, postal codes were retrieved by a specialised data manager.

### 2.2 Imputation

To account for missing data we used multiple imputation. All clinical variables, including outcome, were used to impute missing values [17]. In order to improve imputation results, the Nelson-Aalen estimator for the cumulative hazard was also included as an approximation of the baseline cumulative hazard [18]. The imputation process had 20 iterations and 10 imputations.

### 2.3 Statistical analysis

We determined the median follow-up time using the reverse Kaplan-Meier method [19]. To identify the factors that need to be corrected for, a directed acyclic graph (DAG) was made with all collected clinical variables, using DAGgity v3.0 [20]. The relation between the factors in the DAG was determined with clinical expertise and opinion from an experienced clinician. The DAG allowed us to examine two effects of SES on survival, the total and direct effects. The reason for this distinction is that a determinant can have an effect on the outcome in multiple ways: directly, and through mediating factors. For example, someone’s age might influence the odds of developing cancer. However, age can also have an impact on lifestyle factors (such as smoking or alcohol consumption) which in turn also affects cancer incidence. In other words, lifestyle would be a mediating factor. In this example, the effect of age on cancer incidence directly is the direct effect, and the effect through lifestyle is the indirect effect. Combining the direct and indirect effects gives the total effect, i.e. all possible ways the determinant affects the outcome. Please note that for both direct and total effects, confounding has to be taken into account. With the DAG, you can discover which variables you should correct for to achieve the desired effect: all confounders for the total effect, and all confounders and mediating factors for the direct effect. By looking at and comparing the total and direct effects, it can be concluded whether there was a relation between SES and survival, and whether this association is mostly explained through mediating factors, or only through the direct effect.

We fitted Cox proportional hazards models with SES as a continuous variable, corrected for the variables identified by the DAG to obtain the total and direct effects, and survival time as the outcome. The proportional hazards assumption was assessed for each model by correlating the scaled Schoenfeld residuals with time, confirming their independence. Variables that led to violation of this assumption were entered to the model as strata. Linearity of both SES and mediating factors was assessed by including them as a 5-knot spline, and checking whether this improved model fit with likelihood ratio tests and the Akaike information criterion (AIC).

The SES score was divided into tertiles (in line the instructions provided by the SCP), and the survival in each groups was estimated with Kaplan Meier analysis to obtain median survival, 1-year and 2-year survival rates, and a survival plot. Comparison of the survival difference between the SES tertiles was done with a log-rank test, as well as Cox proportional hazards models corrected to obtain the total and direct effects. To account for possible confounding, an adjusted Kaplan Meier analysis was also estimated balancing the SES tertiles for differences in clinical variables by using inverse probability weighting (IPW). Weights were determined with multinomial regression, reflecting the probability of a subject having their observed SES tertile, given all potential predictive variables [21,22]. We performed this analysis twice, with the variables needed for both the total and the direct effect, in each using age with a 5-knot spline. From each analysis, we obtained a survival plot, median survival, and 1-year and 2-year survival rates.

Both the Cox proportional hazards modelling and Kaplan Meier analysis were done within the individual imputed datasets, and then pooled. For the HR’s, pooling was done in accordance with the Rubin’s rules.[23] For IPW adjustment, the weights from the datasets were averaged for each patient.

We calculated effect estimates with corresponding 95% confidence intervals, and considered a two-sided p-value smaller than 0.05 as statistically significant. Statistical analyses were performed with R 3.5.1 (R Foundation for Statistical Computing, Vienna, Austria) with use of the packages ‘survival’ v3.1-8, ‘mice’ v3.8.0, ‘ipw’ v1.0-11, and ‘IPWsurvival’ v0.5 [24].

## 3. Results

### 3.1 Participants

Between 2012 and 2017, 466 patients were treated with SRS or FSRT for brain metastases. Of those, 404 had one of the five primary tumour types of interest. Median follow-up time was 42.5 months (IQR 29.3

– 67.6), during which 348 (86.1%) patients deceased. The SES ranged from -2.7 to 2.6, with a median of 0.5, and an IQR of 0.1 – 1.0. There were relatively more men in the highest SES tertiles, and NSCLC was more present in the lowest tertile (**Table 1**). Other characteristics were well distributed among the SES groups. Single-fraction SRS was used in 233 patients (57.7%), with the rest receiving FSRT (**Table 2**). Multiple imputation was performed and checked, resulting in a complete dataset of 404 cases. Thirty-eight patients (9.4%) had missing data, of the following variables: NSCLC adenocarcinoma yes or no (25 missings, 6.2%), BRAF mutation (1; 0.2%), breast cancer pathology (2; 0.5%), Hb (9; 2.2%), and SES score (1; 0.2%).

**Table 1:** Baseline characteristics of included patients

|  |  | **Low SES**<br>**[-2.7 – 0.2]**<br>**n=136** | **Mid SES**<br>**[0.2 – 0.8]**<br>**n=133** | **High SES**<br>**[0.8 – 2.6]**<br>**n=134** |
| --- | --- | --- | --- | --- |
| **Age at diagnosis in years, mean (SD)** |  | 62 (10) | 62 (10) | 64 (10) |
| **Sex, n (%)** | Male | 59 (43.4) | 63 (47.4) | 76 (56.7) |
|  | Female | 77 (56.6) | 70 (52.6) | 58 (43.3) |
| **KPS, n (%)** | ≤70 | 49 (36.0) | 38 (28.6) | 46 (34.3) |
|  | 80 | 54 (39.7) | 54 (40.6) | 46 (34.3) |
|  | 90-100 | 33 (24.3) | 41 (30.8) | 42 (31.3) |
| **Tumour type, n (%)** | NSCLC | 86 (63.2) | 79 (59.4) | 70 (52.2) |
|  | Breast | 16 (11.8) | 16 (12.0) | 16 (11.9) |
|  | Melanoma | 11 (8.1) | 12 (9.0) | 12 (9.0) |
|  | RCC | 7 (5.1) | 11 (8.3) | 11 (8.2) |
|  | GI | 16 (11.8) | 15 (11.3) | 25 (18.7) |
| **Extracranial metastasis, n (%)** | Yes | 70 (51.5) | 73 (54.9) | 74 (55.2) |
|  | No | 66 (48.5) | 60 (45.1) | 60 (44.8) |
| **Number of BM, n (%)** | 1 | 68 (50.0) | 69 (51.9) | 65 (48.5) |
|  | 2 | 33 (24.3) | 27 (20.3) | 28 (20.9) |
|  | 3 | 25 (18.4) | 22 (16.5) | 24 (17.9) |
|  | ≥4 | 10 (7.4) | 15 (11.3) | 17 (12.7) |
| **EGFR/ALK status, n (%)\*** | Positive | 6 (7.0) | 9 (11.4) | 4 (5.7) |
|  | Negative | 80 (93.0) | 70 (88.6) | 66 (94.3) |
| **ER/PR status, n (%)\*** | Positive | 10 (62.5) | 4 (25.0) | 6 (37.5) |
|  | Negative | 6 (37.5) | 12 (75) | 10 (62.5) |
| **HER2 status, n (%)\*** | Positive | 8 (50.0) | 8 (50.0) | 10 (62.5) |
|  | Negative | 8 (50.0) | 7 (43.8) | 5 (31.3) |
|  | Missing | - | 1 (6.3) | 1 (6.3) |
| **BRAF status, n (%)\*** | Positive | 5 (45.5) | 6 (50.0) | 4 (33.3) |
|  | Negative | 5 (45.5) | 6 (50.0) | 8 (66.7) |
|  | Missing | 1 (9.1) | - | - |
| **Hb, mean (SD)\*** |  | 8.2 (0.5) | 8.5 (1.1) | 7.8 (0.8) |
| **Missing, n (%)** |  | 4 (57.1) | 3 (27.3) | 2 (18.2) |

BM: brain metastasis, GI: gastro-intestinal, KPS: Karnofsky Performance Status, NSCLC: non-small cell lung carcinoma, RCC: renal cell carcinoma, SD: standard deviation, SES: socioeconomic status
\* Data for relevant primary tumour type

**Table 2.** Delivered radiotherapy doses and fractionation

| Type | Fractions and dose | n (%) |
| --- | --- | --- |
| SRS | 1 x 16 Gy | 6 (1.5%) |
|  | 1 x 18 GY | 55 (13.6%) |
|  | 1 x 20 Gy | 93 (23.0%) |
|  | 1 x 21 Gy | 62 (15.3%) |
|  | 1 x 24 Gy | 17 (4.2%) |
| FSRT | 3 x 8 Gy | 147 (36.4%) |
|  | 5 x 6 Gy | 18 (4.5%) |
| Other | 10 x 30 Gy | 5 (1.2%) |

FSRT = fractionated radiotherapy; SRS = stereotactic radiosurgery

### 3.2 Effect of SES on survival

The entire cohort had a median survival of 10.1 months (95% CI 8.4-11.9), with a 1-year and 2-year survival of 44.6% (95%CI 40.0-49.7) 25.8% (95%CI 21.8-30.4).

Unadjusted, estimation of the effect SES on survival resulted in a HR of 0.92 (95%CI 0.82 – 1.04) for each unit increase in SES score, representing 1 standard deviation within the normal population. The DAG which was used to identify variables for which to correct is shown in **Figure 1**. All collected clinical variables were used in creating the DAG.

**Figure 1:**
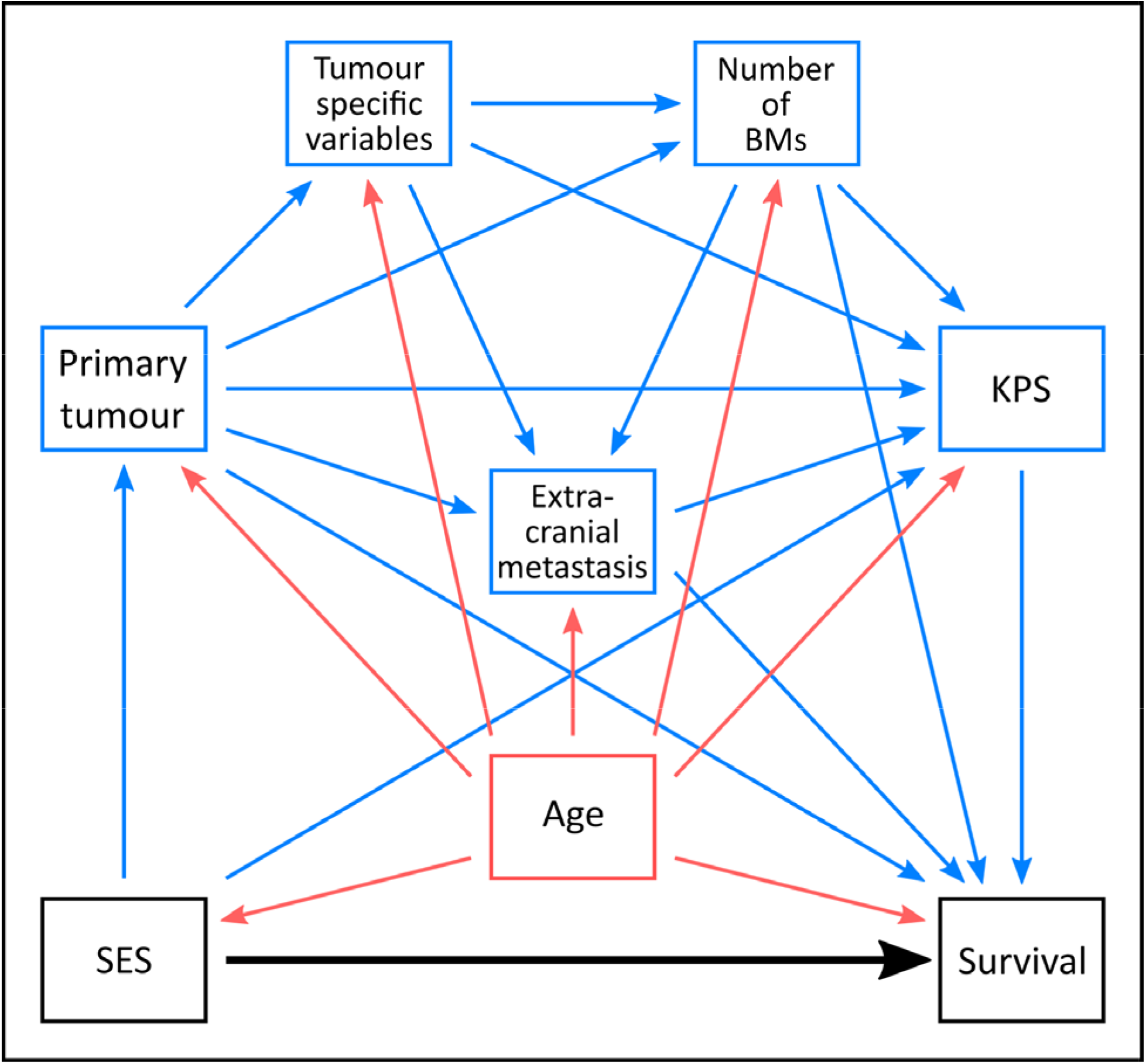
Directed acyclic graph (DAG), showing the relation between all collected clinical variables. Shown are the direct relation between the determinant and outcome (black arrow), and the influence of confounders (red) and mediating factors (blue). BM = brain metastases, KPS = Karnofsky Performance Status, SES = socioeconomic status

In accordance with the DAG, the estimation of total effect of SES on survival was achieved by correcting only for the confounder age, resulting in a HR of 0.95 (95% CI 0.84 – 1.07). The direct effect was determined by correcting for age, extracranial metastasis, KPS, number of brain metastases, and primary tumour location. This gave a HR of 0.96 (95%CI 0.84 – 1.10). None of the estimated HRs for SES were statistically significant within the imputed dataset. All models conformed to assumptions of linearity and proportional hazards, after the appropriate adjustments were made. This included a 5-knot spline for age, and entering KPS and the number of brain metastases as strata to the regression model for the direct effect.

Unadjusted median survival in the low, middle and high tertile was respectively 10.0, 8.8, and 10.5 months (**Table 3**). Adjusting to obtain the direct and total effects resulted in similar survival numbers, and non-significant HRs of 1.1 for middle vs low SES, and 1.0 of high vs low SES. The unadjusted and IPW-adjusted Kaplan Meier survival curves (**Figures 2 and 3**) both show similar survival for each SES tertile. The latter curve was adjusted with the variables needed to obtain the direct effect, the survival curves for the total effect was similar in shape. The log-rank test of the unadjusted Kaplan Meier plot resulted in a non-significant difference, with a chi-square of 0.5 and a p-value of 0.8.

**Table 3.**
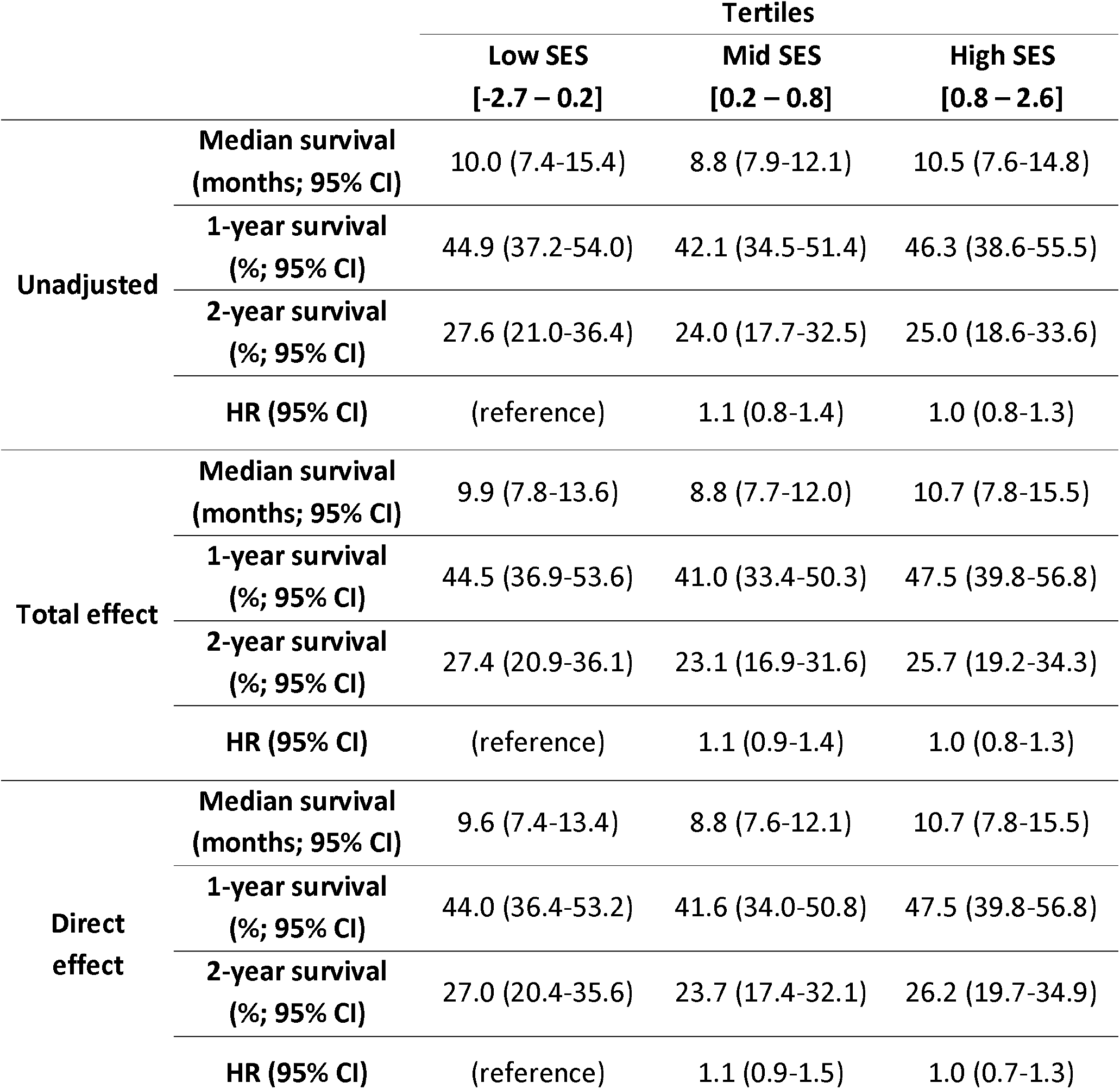
Median survival, survival rates and hazard ratios of each SES tertile, taken from the unadjusted, total and direct effects models

|  |  | Tertiles |  |  |
| --- | --- | --- | --- | --- |
|  |  | Low SES<br>[-2.7 – 0.2] | Mid SES<br>[0.2 – 0.8] | High SES<br>[0.8 – 2.6] |
| Unadjusted | Median survival<br>(months; 95% CI) | 10.0 (7.4-15.4) | 8.8 (7.9-12.1) | 10.5 (7.6-14.8) |
|  | 1-year survival<br>(%; 95% CI) | 44.9 (37.2-54.0) | 42.1 (34.5-51.4) | 46.3 (38.6-55.5) |
|  | 2-year survival<br>(%; 95% CI) | 27.6 (21.0-36.4) | 24.0 (17.7-32.5) | 25.0 (18.6-33.6) |
|  | HR (95% CI) | (reference) | 1.1 (0.8-1.4) | 1.0 (0.8-1.3) |
| Total effect | Median survival<br>(months; 95% CI) | 9.9 (7.8-13.6) | 8.8 (7.7-12.0) | 10.7 (7.8-15.5) |
|  | 1-year survival<br>(%; 95% CI) | 44.5 (36.9-53.6) | 41.0 (33.4-50.3) | 47.5 (39.8-56.8) |
|  | 2-year survival<br>(%; 95% CI) | 27.4 (20.9-36.1) | 23.1 (16.9-31.6) | 25.7 (19.2-34.3) |
|  | HR (95% CI) | (reference) | 1.1 (0.9-1.4) | 1.0 (0.8-1.3) |
| Direct effect | Median survival<br>(months; 95% CI) | 9.6 (7.4-13.4) | 8.8 (7.6-12.1) | 10.7 (7.8-15.5) |
|  | 1-year survival<br>(%; 95% CI) | 44.0 (36.4-53.2) | 41.6 (34.0-50.8) | 47.5 (39.8-56.8) |
|  | 2-year survival<br>(%; 95% CI) | 27.0 (20.4-35.6) | 23.7 (17.4-32.1) | 26.2 (19.7-34.9) |
|  | HR (95% CI) | (reference) | 1.1 (0.9-1.5) | 1.0 (0.7-1.3) |

**Figure 2:**
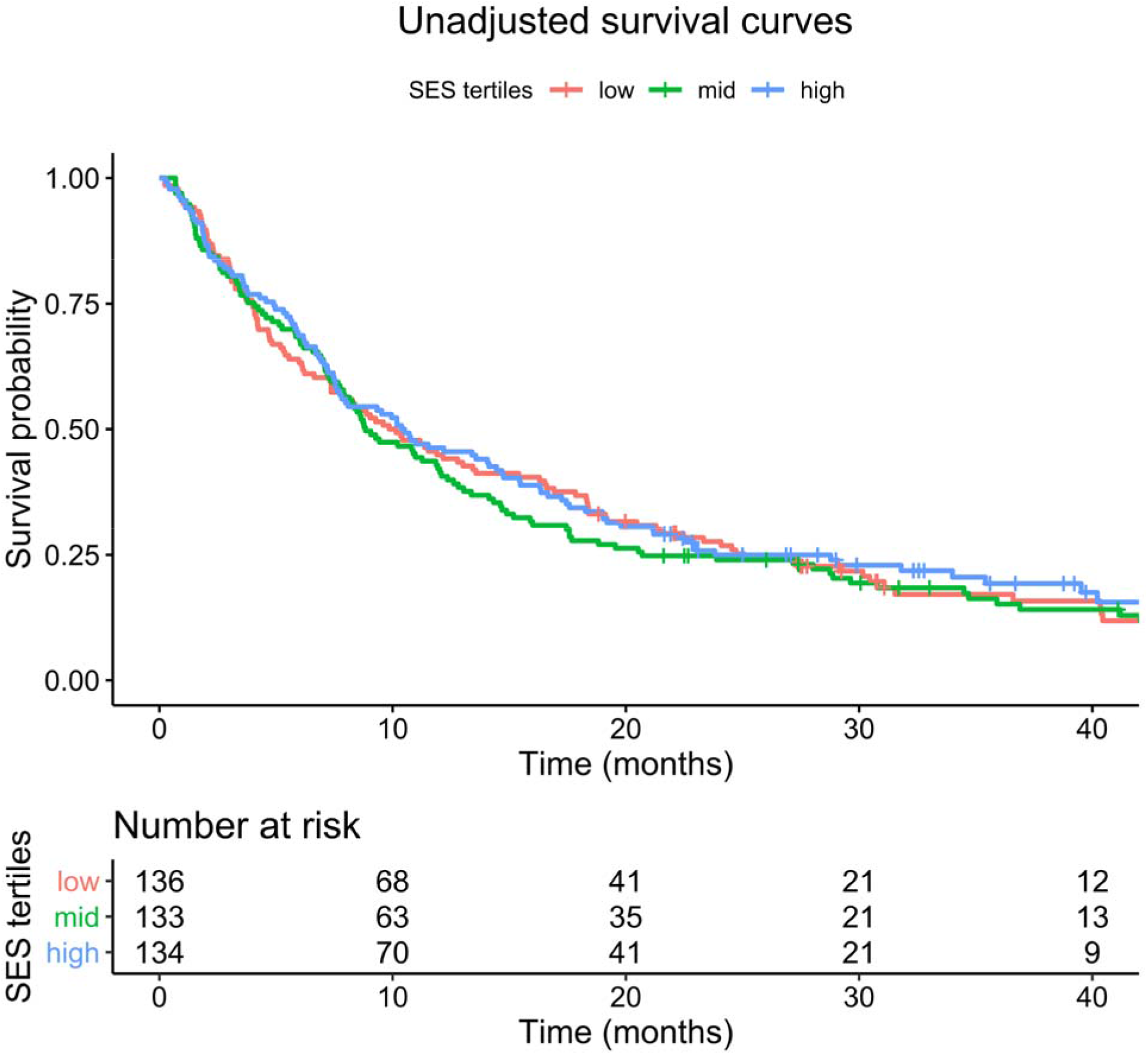
Survival curves comparing the SES tertiles, unadjusted for other clinical variables (logrank test p=0.8)

**Figure 3:**
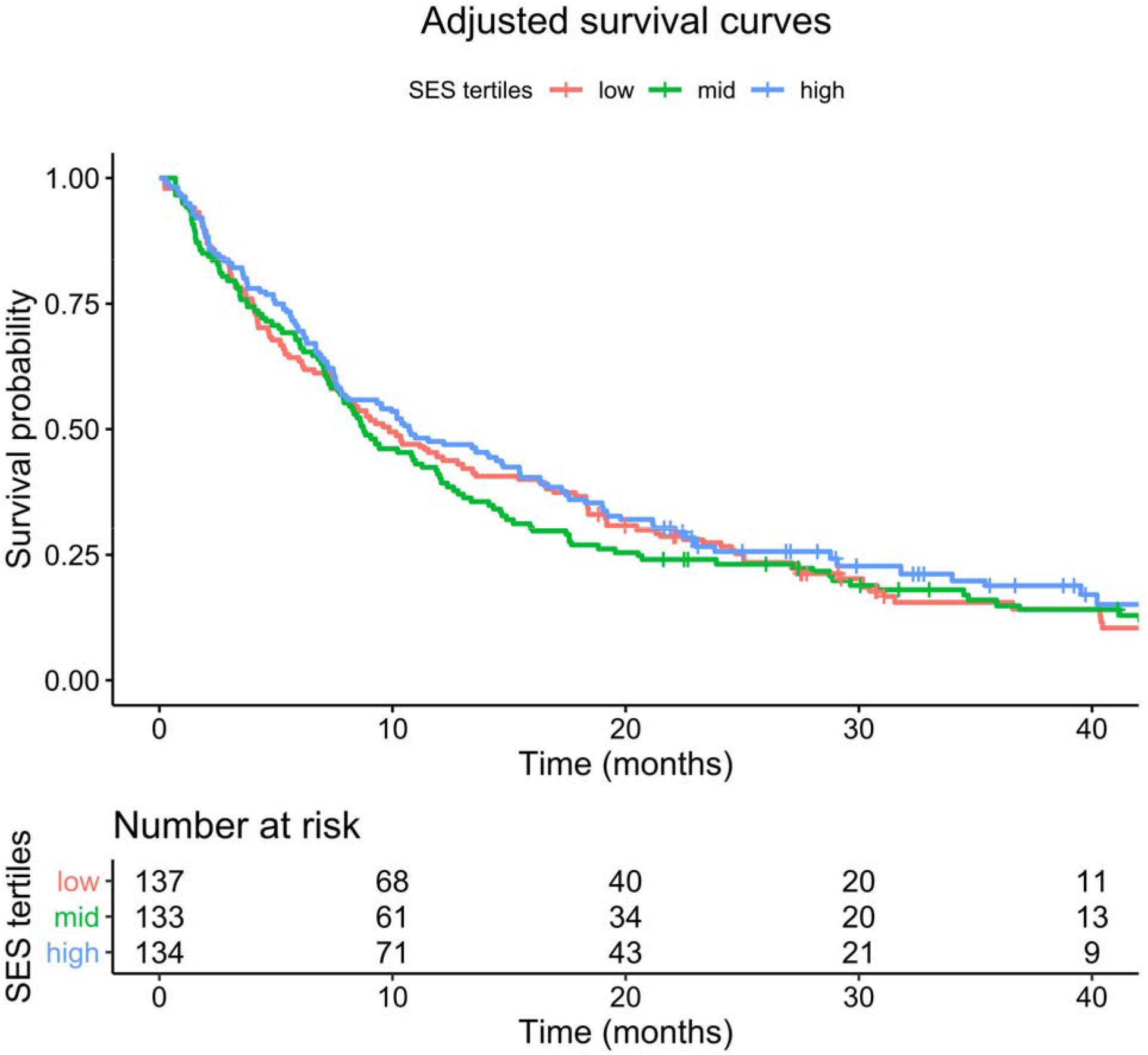
Survival curves comparing the SES tertiles, adjusted with inverse probability weighting for the clinical variables needed to obtain the direct effect

## 4. Discussion

We have found that socioeconomic status (SES) is not related to overall survival in patients with brain metastases referred for and treated with stereotactic radiosurgery in a tertiary radiotherapy facility in the Netherlands. Both in unadjusted analysis as well as when adjusting for other clinical factors, we found a non-significant relation between SES and survival, with HRs and confidence intervals suggesting no relevant clinical impact either. The survival curves lead to a similar conclusion, showing no clear differences in survival patterns between the SES tertiles.

This not the first study into the relation between SES and survival in brain metastasis patients. Previous studies have been performed, with differing results.

Ten Berge et al studied a cohort of 1129 Dutch patients with synchronously diagnosed brain metastases and NSCLC treated with Gamma Knife (GK) radiosurgery [15]. They found a significant relation between SES (in tertiles, based on the same SCP database as the present study) and survival, with HRs of 0.9 (95% CI 0.7-0.995) and 0.8 (95% CI 0.7-0.9) for the intermediate and high SES groups, compared to the low group. Analysis was corrected for a large number of factors, including age, gender, and primary tumour stage and adenocarcinoma histology. Their observed point estimates are close to the HRs we found (0.95 and 0.96), and thereby are close to 1. This could lead one to argue that, even though there is a significant effect, its clinical impact is limited. The authors themselves did not reflect on this effect of SES on survival, as their primary aim was to study whether there are factors that influence referral patterns in these patients. They found that patients had increased odds to be referred for GK if they were below 60 years of age and had limited lymphatic spread (i.e. lower N-stage). This (aside from the more selective study population) might partly explain the difference to our results, although SES did not affect the referral pattern.

A cohort of 737 patients undergoing SRS for brain metastases from the most common primary tumour sites in the United States was studied by Alphonse-Sullivan et al. [12]. They examined four factors reflecting a patient’s SES: median income (based on postal code), race, rural or urban residence, and health insurance status. They found a significant result for median income, but mention there is no increasing or decreasing trend across the income groups. None of the other factors was associated with survival. The absence of income data in our cohort means we cannot compare these results to ours.

A Chinese study in 209 NSCLC patients with multiple brain metastases found a significant effect of a patient’s “cultural background” on survival [14]. This metric was composed of socioeconomic status, level of education, understanding of the disease, and the degree of care and support they received from family members. It is unclear how these factors were measured or defined, meaning our results cannot be compared to theirs. They concluded that, compared to the highest level, the two lowest cultural background groups had significantly poorer survival.

Finally, two studies examined the effect of SES of cohorts including, but not limited to, brain metastasis patients. They did not find any relation between SES and survival [13,25].

Survival is not the only outcome of interest in studying the effect of SES in brain metastasis patients. Along with the previously mentioned study by Ten Berge et al., two other recent studies have looked at the effect of SES on treatment decisions [26,27], which may – in turn – impact survival for the total population of patients with brain metastases. The first study found no relation between referral patterns, but the latter two have found that patients within a higher SES quartile are more likely to receive SRS.

There are some limitations to this study. Firstly, this is relatively small cohort, especially compared to that of Ten Berge et al. This might mean that our sample size is too small to find a significant relationship between SES and survival. However, the HRs found both in our study and that by Ten Berge et al. suggest limited clinical relevance, even if a significant effect is present.

Secondly, our SES score was based on a patients postal code, meaning that there may be imprecisions in this metric. Should a patient be an outlier in terms of education, income or unemployment, this would not be reflected their assigned SES score. This means the SES data we used for analysis can be either too high or too low, but is always based on an average score of a patients’ neighbourhood. This could have led to an underestimation of the observed effect. There is no consensus as to which way of determining SES is the best in cancer-related survival studies [28], which may also explain the variance in previously published studies. Different methods may result in contrasting conclusions, meaning the SES scoring type should be chosen carefully. Unfortunately, the currently used SES score is the only one available to us, so a comparison between different scoring metrics cannot be performed. However, we have used the same SES scores as Ten Berge et al., which increases comparability of the outcomes.

As with most retrospective studies, we were faced with missing data. Since the rate of missing data was relatively low, and as we used multiple imputation (currently the best method available to minimize bias), we feel that the effects of these missing data was limited.

Finally, and perhaps most importantly, selection bias plays a role. We have selected only those patients receiving SRS or FSRT for the 5 most common primary sites. This excludes a number of brain metastases patients, particularly those receiving whole brain radiotherapy, or those not receiving radiation treatment at all. This means that we do not have a representative cohort that allows us to study the effect of SES of the entire brain metastasis population, but only on the SRS/FSRT-treated subgroup. As mentioned above, referral patterns for SRS can be affected by SES, which in turn has an effect on survival. This means that treatment referral may be an additional mediator between SES and survival, but one we cannot correct for.

Our results warrant several directions for future studies. In order to conclusively say that SES does not affect survival, and in light of our major two limitations, a bigger cohort of brain metastasis patients should be selected for whom all relevant SES factors are recorded. As this includes annual income and employment status, which is not routinely recorded in clinical practice, this would require prospective data collection. This cohort should also include those patients who did not receive SRS or FSRT, to reflect the entire brain metastasis population. Additionally, we have seen that difference in local or national healthcare policies might influence differences seen between SES groups. A large-scale cohort comprised of several countries representing different healthcare systems (e.g. nationalized vs private healthcare, obligatory health insurance yes or no) could lead to conclusions on the level of equality of treatment outcomes in these systems.

## Conclusion

We have studied the effect of socioeconomic status on survival in a cohort of brain metastases patients receiving either SRS or FSRT. When correcting for clinical variables, we found no significant relationship between the estimated SES and survival, with HRs suggesting limited clinical impact. Although our study has certain limitations, including selection bias due to treatment referral patterns, the results suggest that patients’ survival outcomes after contemporary cancer treatment are unrelated to their employment status, education status and annual income.

## Data Availability

All data produced in the present study are available upon reasonable request to the authors

## Acknowledgments

Special thanks to Gery Tijsseling for providing data management for this study.

## Abbreviations

AIC: Akaike information criterion
DAG: directed acyclic graph
DS-GPA: disease-specific Graded Prognostic Assessment
GBA: *Gemeentelijke Basisadministratie* (Municipal Personal Records Database)
GI: gastro-intestinal
GK: Gamma Knife
IPW: inverse probability weighting
KPS: Karnofsky performance status
NSCLC: non-small-cell lung carcinoma
RCC: renal cell cancer
RT: radiotherapy
SCP: (Netherlands Institute for Social Research)
SES: socioeconomic status
SRS: stereotactic radiosurgery
FSRT: fractionated stereotactic radiotherapy

